# White Matter Hyperintensity Burden Modifies the Association Between Atrial Fibrillation and Cerebral Microbleeds

**DOI:** 10.64898/2026.06.03.26354875

**Authors:** Wi-Sun Ryu, Leonard Sunwoo, Myungjae Lee, Kyusik Kang, Jae Guk Kim, Soo Joo Lee, Jae-Kwan Cha, Tai Hwan Park, Jeong-Yoon Lee, Kyungbok Lee, Doo Hyuk Kwon, Jun Lee, Hong-Kyun Park, Yong-Jin Cho, Keun-Sik Hong, Minwoo Lee, Mi Sun Oh, Kyung-Ho Yu, Dong-Seok Gwak, Dong-Eog Kim, Hyunsoo Kim, Joon-Tae Kim, Joong-Goo Kim, Jay Chol Choi, Wook-Joo Kim, Jee Hyun Kwon, Kyu Sun Yum, Dong-Ick Shin, Jeong-Ho Hong, Sung-Il Sohn, Sang-Hwa Lee, Chulho Kim, Hae-Bong Jeong, Kwang-Yeol Park, Keon-Joo Lee, Chi Kyung Kim, Jihoon Kang, Jun Yup Kim, Hee-Joon Bae, Beom Joon Kim

## Abstract

**Background:** In atrial fibrillation (AF), cerebral microbleed (CMB) burden guides anticoagulation decisions, yet AF is itself inconsistently associated with CMBs, a paradox unexplained by frameworks that treat CMBs as a unitary marker of small vessel disease. We hypothesized that the white matter hyperintensity (WMH) context in which CMBs arise modifies their vascular meaning, and that this context-dependence underlies the inconsistent AF–CMB association.

**Methods:** From a multicenter Korean stroke registry, we analyzed 5,735 first-ever ischemic stroke patients imaged at nine centers using susceptibility-weighted MRI. WMH volume and CMB count were extracted by validated deep learning pipelines. Patients were cross-classified by age-adjusted WMH residual (median split) and CMB count (≥2) into four groups. The AF–CMB association was estimated by multivariable logistic regression within each WMH stratum with formal interaction testing. Spatial CMB distribution was analyzed against the Automated Anatomical Labeling atlas.

**Results:** In the full cohort (mean age 69.5 years; 57.7% male), AF was not associated with CMBs (OR 1.04; 95% CI 0.87–1.25). Stratification yielded divergent estimates: the adjusted AF OR was 1.46 (1.11–1.93; P = 0.007) in the WMH-low stratum and 0.95 (0.73–1.22; P = 0.665) in the WMH-high stratum, with significant interaction (OR 0.56; P < 0.001). The discordant phenotype (low WMH, high CMB; 8.9%) was enriched for AF (28.0%) and showed fronto-temporal cortical predominance with deep structure sparing. AF independently reduced the proportion of deep CMBs (IRR 0.80; P = 0.040). The interaction was preserved across prespecified sensitivity analyses.

**Conclusions:** The AF–CMB association is confined to patients with low WMH burden relative to age and is accompanied by a topographically distinct CMB distribution. Clinical assessment of small vessel disease based on WMH alone may overlook a CMB phenotype linked to AF.

## Introduction

Anticoagulation reduces ischemic stroke recurrence in patients with atrial fibrillation, yet the prescribing decision is increasingly conditioned by the presence of cerebral microbleeds (CMBs), which mark hemorrhage-prone microvasculature and rank among the strongest imaging predictors of intracranial hemorrhage in anticoagulated patients.^1–4^ Clinical interpretation of CMBs rests on a topographic schema that ascribes lobar lesions to cerebral amyloid angiopathy and deep or infratentorial lesions to arteriolosclerotic small vessel disease.^5,6^ White matter hyperintensity (WMH) volume is the principal aggregate marker of small vessel injury,^7–11^ and where WMH and CMB burdens track together, this dual framework supplies a coherent account. A substantial subset of patients, however, departs from concordance: their CMB burden is disproportionate to their WMH load, and the existing categories are silent on what such a discrepancy represents.

Whether this discordance reflects measurement variability, the differential age sensitivity of the two markers, or a genuinely distinct vascular phenotype has not been resolved. Prior investigations have either summed WMH and CMB into composite small vessel disease scores or examined them independently, neither of which can determine whether their relative imbalance carries pathophysiological meaning.^4^ The pertinent question is not whether discordant patients exist, since they plainly do, but whether the discordance itself indexes a vascular biology that conventional small vessel disease frameworks were not constructed to detect.

Two methodological obstacles have rendered this question difficult to address. First, both WMH volume and CMB count rise steeply with age,^12–15^ so a naive cross-classification of the two markers largely recapitulates an age stratification rather than isolating phenotypic divergence. Second, identifying a small subgroup defined by marker discordance demands quantitative imaging measurements applied uniformly across a cohort large enough to retain statistical power within each cell of the cross-classification, a combination not previously available at scale. Recent advances in deep learning–based segmentation now permit uniform extraction of WMH volume and CMB count across thousands of clinical MRI studies,^16^ and an age-adjusted residual definition of WMH burden preserves the WMH axis while removing its confounding with age.

In 5,735 first-ever ischemic stroke patients drawn from a nationwide multicenter registry,^17,18^ we therefore sought to determine whether patients with discordant WMH and CMB burdens constitute a clinically and pathophysiologically distinct phenotype. We characterized their risk factor profiles, stroke etiologies, and one-year outcomes; tested whether the previously inconsistent association between atrial fibrillation and CMBs^19,20^ differs across strata of age-adjusted WMH burden; and analyzed the spatial distribution of CMBs to determine whether the topographic signature of the discordant phenotype conforms to, or departs from, the patterns predicted by amyloid angiopathy and arteriolosclerosis.

## Materials and methods

### Study population

The CRCS-K (Clinical Research Collaboration for Stroke in Korea) image repository is a multicenter archive linked to a prospective nationwide stroke registry, with standardized clinical, laboratory, imaging, and outcome data collected through a structured web-based platform with periodic quality audits.^17,18^ Because accurate CMB ascertainment requires hemorrhage-sensitive imaging, the analysis was restricted to nine centers where susceptibility-weighted imaging (SWI) was the dominant gradient-echo sequence (adoption rate > 80%). SWI detects substantially more hemosiderin deposits than gradient-recalled echo T2*,^21–23^ so only patients imaged with SWI were retained.

Within the nine SWI-dominant centers, inclusion required a confirmed diagnosis of acute ischemic stroke by diffusion-weighted imaging (DWI), with available fluid-attenuated inversion recovery (FLAIR) and SWI sequences adequate for quantitative extraction of WMH volume, lacune count, and CMB count. Patients were excluded if they had a history of prior stroke, to avoid confounding by antecedent cerebrovascular injury, or if they had received intravenous thrombolysis or endovascular thrombectomy, because hemorrhagic transformation may be misclassified as pre-existing CMBs (Supplementary Fig. 1).

The study was approved by the institutional ethics committees of all participating centers (lead IRB No. B-2308-845-302), with written informed consent obtained from all patients or their representatives in accordance with the Declaration of Helsinki.

### MRI acquisition

All participating centers acquired brain MRI during the acute hospitalization using 1.5T or 3T scanners from three vendors (Siemens [n = 2,788], Philips [n = 2,524], GE [n = 422]; manufacturer information was missing for one patient). Axial FLAIR images were obtained for WMH assessment (TR 3,860–13,000 ms; TE 68–448 ms; slice thickness 3–6 mm, median 5 mm). Axial SWI was acquired for CMB detection (TR 11–84 ms; TE 35–55 ms; slice thickness 1.0–3.0 mm, median 2.0 mm).

### Quantitative MRI marker extraction

WMH volume was segmented on FLAIR images using a validated deep learning algorithm (Dice 0.722; r = 0.933 against manual delineation; Supplementary Methods).^16^ Total brain volume was extracted from the same framework.^16^

CMBs were detected on SWI using a 3D nnU-Net^24^ trained on a multi-institutional dataset of 932 subjects, with sequential post-processing for anatomy-based mimic removal, geometric constraints, and vessel tracking (Supplementary Methods). On external validation, the model achieved subject-level sensitivity of 91.3% and specificity of 84.2%. Native-space and Montreal Neurological Institute (MNI)-registered CMB counts agreed closely (r = 0.966); native-space counts were used for analysis.

Lacunes were identified on FLAIR images using a detection algorithm trained to recognize small (3 to 15 mm), well-demarcated, cerebrospinal fluid-intensity cavities in the white matter, basal ganglia, or brainstem, consistent with the STRIVE-2 criteria for lacunes of presumed vascular origin.^25^

### Variable definitions and group assignment

WMH volume increases strongly with age (R² = 0.31), and a raw volume median split would separate patients into a young low-WMH group and an old high-WMH group (mean age difference 13.5 years), effectively confounding the WMH axis with age. To avoid this, we operationalized the WMH axis as an age-adjusted residual: raw volume was log-transformed (log(volume + 1)) and regressed on age, and the residual from this model was used to classify each patient as WMH-low or WMH-high relative to the median. This approach reduced the between-stratum age difference to 0.7 years (Supplementary Fig. 2).

The CMB axis was dichotomized at a count of two or more as the primary threshold to minimize false-positive classification, since solitary hypointense foci on SWI may represent calcification, vessel cross-sections, or partial-volume artifacts rather than true hemosiderin deposits. Thresholds of one or more and five or more were examined in sensitivity analyses.

Cross-classification of the two axes generated four groups: Group 1 (WMH-low, CMB-low; reference), Group 2 (WMH-low, CMB-high; the discordant group), Group 3 (WMH-high, CMB-low), and Group 4 (WMH-high, CMB-high). AF was ascertained from the registry’s structured fields, which record a documented history of AF or new-onset AF detected during the index hospitalization. Stroke etiology was classified by the treating physician according to the MRI-based modified Trial of ORG 10172 in Acute Stroke Treatment (TOAST) criteria.^26,27^

Clinical outcomes were ascertained through the registry’s structured follow-up at 3 months and 1 year, with stroke recurrence classified as ischemic or hemorrhagic. Cumulative one-year events were defined as the union of events recorded at either time point. Follow-up was available for 5,720 patients (99.7%).

### Microbleed spatial analysis

In patients with at least one CMB (n = 2,474), individual CMB segmentation masks were registered to MNI standard space and overlaid on the Automated Anatomical Labeling (AAL) atlas^28^ to assign each lesion to one of 116 parcellated regions, aggregated into six compartments: frontal, parietal, temporal, occipital, deep (caudate, putamen, pallidum, thalamus), and infratentorial (cerebellum, vermis). CMBs whose centroids fell outside atlas-defined regions (5.9%) were excluded from compartment-level analyses. Patients with at least two anatomically classified CMBs were categorized by the modified Boston criteria as strictly lobar (all classified CMBs in cortical compartments), mixed, or strictly deep/infratentorial. Group-level voxel-wise probability maps were generated by averaging the binary microbleed masks within each group. To compare spatial distribution between Group 2 and Group 4, mean CMB counts per AAL-defined compartment were computed across groups, and a voxel-wise t-statistic map was generated by permutation testing (5,000 permutations) with family-wise error correction. Region-level differences in burden-normalized per-CMB proportions (regional count divided by total CMB count per patient) were tested with a 5,000-iteration patient-level bootstrap and Benjamini–Hochberg false-discovery-rate correction across 59 bilateral AAL regions (details in Figure 3 legend). The top 12 regions ranked by absolute proportion difference are presented in Figure 3c.

To test whether AF shifts CMB distribution independently of total burden, negative binomial regression was fitted with compartment-specific CMB count as the outcome and log-transformed total count as offset, so that the AF incidence rate ratio captures the proportional excess or deficit relative to expected burden. Models adjusted for the same covariates as the primary analysis were fitted in the full spatial cohort, with an AF × WMH-high interaction term.

### Statistical analysis

Baseline characteristics were compared across groups using chi-square or Kruskal–Wallis tests. The dose-response relationship between CMB count (0, 1, 2–4, 5–9, ≥10) and AF prevalence was examined within each WMH stratum, with trend assessed by logistic regression treating CMB count as a continuous predictor. Because both AF and hypertension prevalence are confounded by age, trends were additionally estimated using age-adjusted marginal prevalence at the mean cohort age. The independent association of AF with CMB burden (≥2) was estimated by multivariable logistic regression in the full cohort and within each WMH stratum, adjusted for age, sex, hypertension, diabetes mellitus, dyslipidemia, current smoking, coronary heart disease, and admission National Institutes of Health Stroke Scale (NIHSS) score. Effect modification was tested by a multiplicative interaction term (AF × WMH stratum) in the full-cohort model.

Three supplementary analyses addressed specific alternative explanations. First, pre-admission oral anticoagulant use was examined as a predictor of CMB burden and distribution among AF patients, and as an additional covariate in the primary model, to assess whether the AF–CMB association reflects anticoagulant-related hemorrhage. Second, AF was reclassified as a three-level variable (no AF, new-onset, or prior) to test whether exposure duration modifies the association. Third, age-adjusted WMH residuals were compared between AF-positive and AF-negative patients to evaluate potential collider bias in WMH-group assignment.

Twelve prespecified sensitivity analyses assessed robustness, examining alternative CMB thresholds (≥1 and ≥5), three alternative WMH definitions (raw volume median split, and two tertile-based splits), exclusion of cardioembolic strokes, age-group stratification (three subgroups), additional lacune adjustment, generalized estimating equations for within-site clustering, and additional adjustment for total brain volume to address potential confounding by global brain atrophy. The interaction was additionally fitted with WMH burden as a continuous covariate, modeling AF, the age-adjusted residual, and their multiplicative product within the same covariate set.

The E-value quantified the minimum unmeasured-confounder strength required to explain away the observed effect.^29^ Hemispheric laterality analysis was performed among Group 2 AF-positive patients with unilateral supratentorial DWI lesions, comparing a laterality index ((CMBipsi − CMBcontra)/(CMBipsi + CMBcontra)) and territorial co-localization against zero and against AF-negative patients (Supplementary Methods). This study was conducted and reported in accordance with the STROBE statement. Model diagnostics (Hosmer-Lemeshow goodness-of-fit, variance inflation factors, overdispersion checks, likelihood-ratio tests for interaction) and generalized estimating equations with robust standard errors for site-clustering analyses are detailed in Supplementary Methods. All analyses used Python 3.10 (statsmodels, scipy); two-sided P < 0.05 was considered significant.

Clinical outcomes shown in Figure 4 were analyzed as follows. Adjusted odds ratios for one-year and three-month hemorrhagic stroke recurrence (rare endpoints with quasi-separation in the four-group cross-classification) were estimated by Firth penalized logistic regression. Adjusted odds ratios for poor three-month and one-year functional outcome (modified Rankin Scale [mRS] ≥ 3) used standard logistic regression, and the cumulative odds ratio for ordinal three-month mRS was obtained from a proportional-odds logistic model with a Brant-style score test. All outcome models adjusted for the same covariates as the primary AF–CMB analysis and used Group 1 as the reference. Time-resolved hemorrhagic recurrence (Fig. 4b, c) was estimated by the Aalen–Johansen cumulative incidence function with death from any cause as the competing event (n = 5,633; daily resolution over 12 months from the SWI index date). Group-specific 1-year cumulative incidences are reported with 95% logit-transformed confidence intervals. Cause-specific and Fine–Gray subdistribution hazard ratios were also computed to confirm that the elevated Group 2 signal was not driven by differential mortality (Supplementary Methods).

## Results

### Cohort assembly and group definition

Among 9,926 patients with acute ischemic stroke at the nine stroke centers, 8,624 had a complete small vessel disease imaging panel (WMH volume on FLAIR and CMB count on SWI). Exclusion of 1,673 patients with prior stroke and 1,216 who received intravenous thrombolysis or endovascular thrombectomy yielded a final analytic cohort of 5,735 (Supplementary Fig. 1). The 4,191 excluded patients were older, had higher admission NIHSS (median 5 vs 2), and had a higher prevalence of AF (28.6% vs 15.7%; Supplementary Table 1). The age-adjusted WMH residual reduced the between-group age difference from 13.5 years under a raw volume median split (mean 62.8 vs 76.3 years) to 0.7 years (69.2 vs 69.9 years; Supplementary Fig. 2). Cross-classification at the median residual and a CMB threshold of two or more assigned 2,357 patients (41.1%) to Group 1 (WMH-low/CMB-low), 510 (8.9%) to Group 2 (WMH-low/CMB-high), 1,717 (29.9%) to Group 3 (WMH-high/CMB-low), and 1,151 (20.1%) to Group 4 (WMH-high/CMB-high; Fig. 1a). The remaining age variation across the four groups (Table 1) reflects the CMB axis: because CMB prevalence rises with age, Groups 2 and 4 (mean 74.8 and 73.3 years) were older than Groups 1 and 3 (68.0 and 67.7 years).

**Figure 1.**
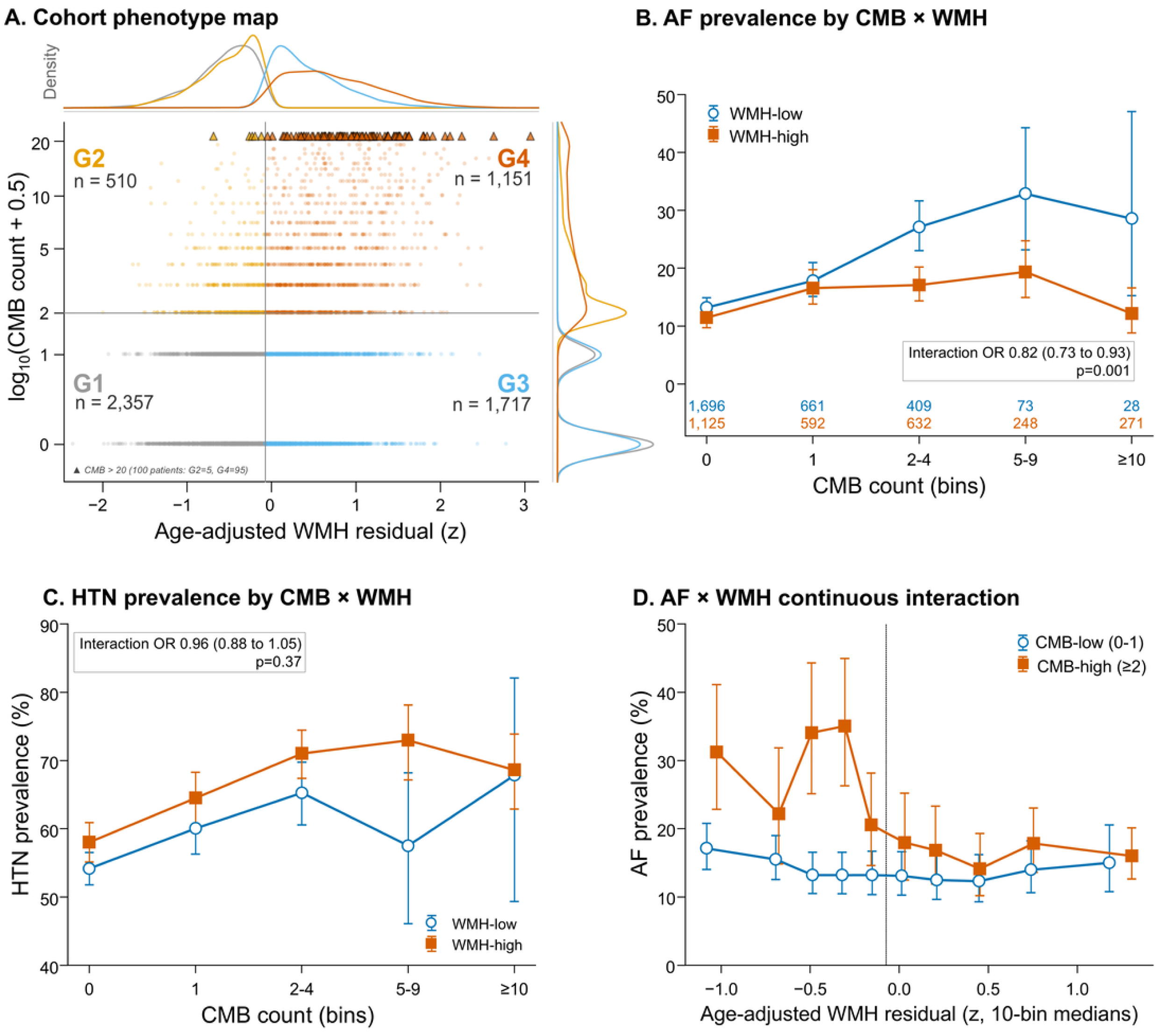
Atrial fibrillation associates with cerebral microbleed burden only in the white matter hyperintensity–low stratum. (a) Cohort phenotype map of the 5,735 first-ever ischemic stroke patients (excluding those who received intravenous thrombolysis or endovascular thrombectomy). Position encodes age-adjusted white matter hyperintensity residual (x-axis) and log10(microbleed count + 0.5) (y-axis); color encodes the four phenotypic groups. Box annotations give per-group N, median age, and atrial fibrillation and hypertension prevalence. Dashed lines mark the group-defining cutoffs. (b) Atrial fibrillation prevalence by microbleed count categories (0, 1, 2–4, 5–9, ≥10), stratified by white matter hyperintensity burden (median split). Open circles, white matter hyperintensity–low; filled squares, white matter hyperintensity–high. Error bars are 95% Wilson confidence intervals; sample size per cell printed below the x-axis. (c) Hypertension prevalence by the same microbleed × white matter hyperintensity grid serves as a negative control. (d) Continuous interaction: patients with microbleed count 0–1 (open circles) and ≥ 2 (filled squares) are binned into 10 quantiles of age-adjusted white matter hyperintensity residual, with atrial fibrillation prevalence computed per bin (95% Wilson confidence intervals). The interaction OR shown in panel (b) reflects the trend interaction (CMB count as continuous ordinal × WMH-stratum binary) on AF prevalence and the panel (d) annotation reflects the CMB-high × WMH-residual continuous interaction; both differ from the primary multiplicative interaction reported in Table 2 (CMB ≥ 2 × WMH-stratum binary, OR 0.56). *Alt text*: Four-panel composite. Panel a is a scatter plot of cohort phenotype with age-adjusted white matter hyperintensity residual on the x-axis and log-transformed microbleed count on the y-axis, color-coded into four groups by quadrant. Panel b is a line graph showing atrial fibrillation prevalence rising with microbleed count only in the white matter hyperintensity-low stratum. Panel c shows hypertension prevalence as a flat negative-control comparison. Panel d is a binned plot showing atrial fibrillation odds declining monotonically across white matter hyperintensity quantiles in patients with at least two microbleeds.

**Table 1.**
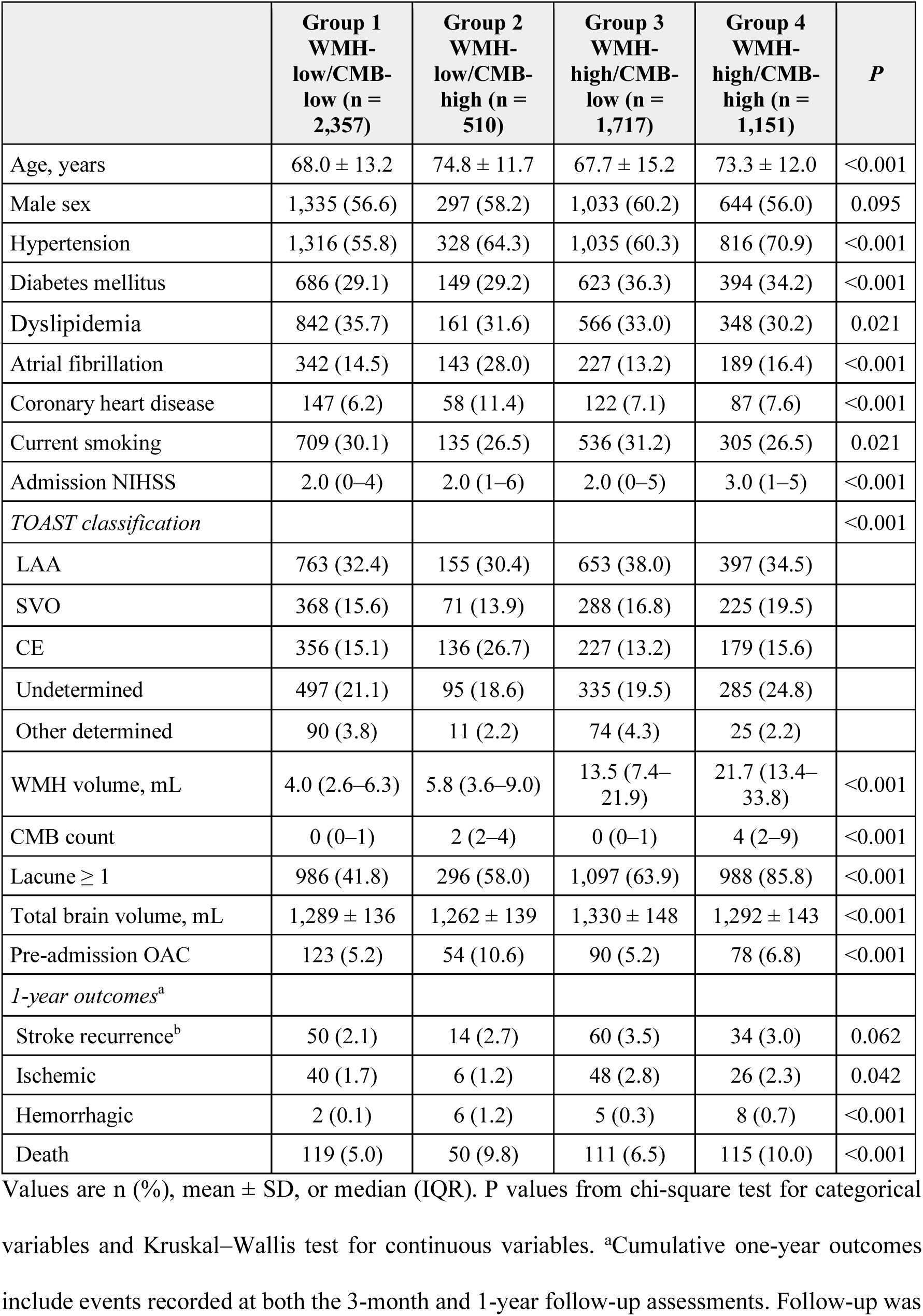
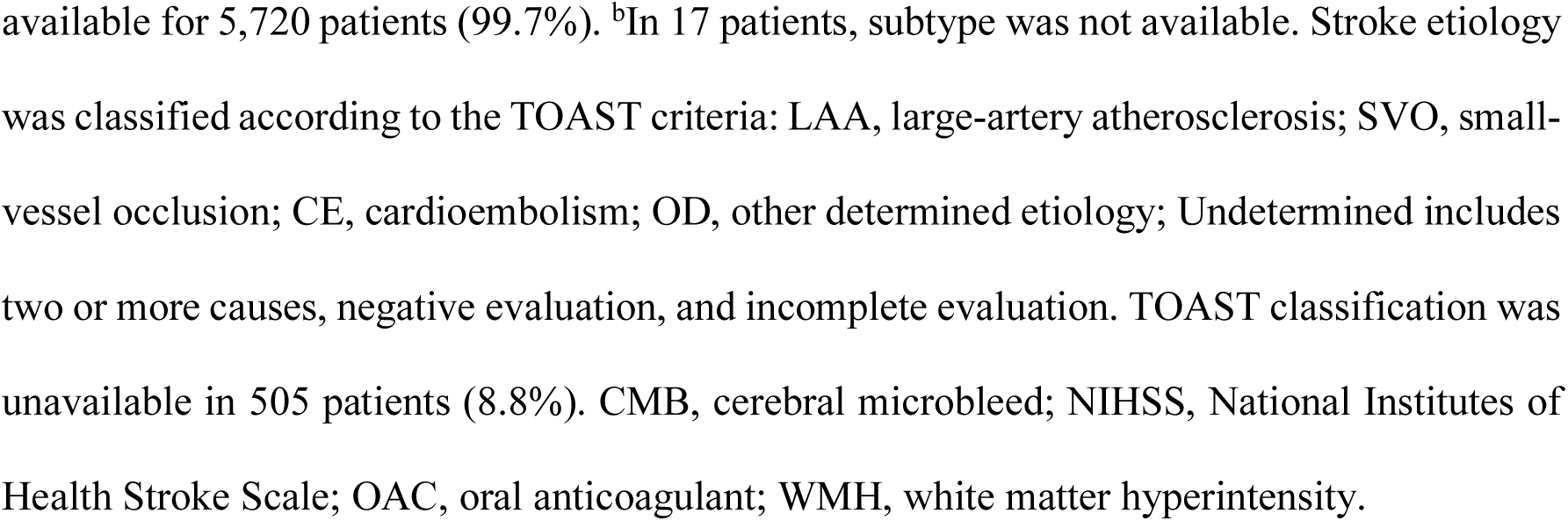
Baseline characteristics of the study population by WMH–CMB group.

|  | **Group 1**<br>**WMH-**<br>**low/CMB-**<br>**low (n =**<br>**2,357)** | **Group 2**<br>**WMH-**<br>**low/CMB-**<br>**high (n =**<br>**510)** | **Group 3**<br>**WMH-**<br>**high/CMB-**<br>**low (n =**<br>**1,717)** | **Group 4**<br>**WMH-**<br>**high/CMB-**<br>**high (n =**<br>**1,151)** | ***P*** |
| --- | --- | --- | --- | --- | --- |
| Age, years | 68.0 ± 13.2 | 74.8 ± 11.7 | 67.7 ± 15.2 | 73.3 ± 12.0 | <0.001 |
| Male sex | 1,335 (56.6) | 297 (58.2) | 1,033 (60.2) | 644 (56.0) | 0.095 |
| Hypertension | 1,316 (55.8) | 328 (64.3) | 1,035 (60.3) | 816 (70.9) | <0.001 |
| Diabetes mellitus | 686 (29.1) | 149 (29.2) | 623 (36.3) | 394 (34.2) | <0.001 |
| Dyslipidemia | 842 (35.7) | 161 (31.6) | 566 (33.0) | 348 (30.2) | 0.021 |
| Atrial fibrillation | 342 (14.5) | 143 (28.0) | 227 (13.2) | 189 (16.4) | <0.001 |
| Coronary heart disease | 147 (6.2) | 58 (11.4) | 122 (7.1) | 87 (7.6) | <0.001 |
| Current smoking | 709 (30.1) | 135 (26.5) | 536 (31.2) | 305 (26.5) | 0.021 |
| Admission NIHSS | 2.0 (0–4) | 2.0 (1–6) | 2.0 (0–5) | 3.0 (1–5) | <0.001 |
| *TOAST classification* |  |  |  |  | <0.001 |
| LAA | 763 (32.4) | 155 (30.4) | 653 (38.0) | 397 (34.5) |  |
| SVO | 368 (15.6) | 71 (13.9) | 288 (16.8) | 225 (19.5) |  |
| CE | 356 (15.1) | 136 (26.7) | 227 (13.2) | 179 (15.6) |  |
| Undetermined | 497 (21.1) | 95 (18.6) | 335 (19.5) | 285 (24.8) |  |
| Other determined | 90 (3.8) | 11 (2.2) | 74 (4.3) | 25 (2.2) |  |
| WMH volume, mL | 4.0 (2.6–6.3) | 5.8 (3.6–9.0) | 13.5 (7.4–21.9) | 21.7 (13.4–33.8) | <0.001 |
| CMB count | 0 (0–1) | 2 (2–4) | 0 (0–1) | 4 (2–9) | <0.001 |
| Lacune ≥ 1 | 986 (41.8) | 296 (58.0) | 1,097 (63.9) | 988 (85.8) | <0.001 |
| Total brain volume, mL | 1,289 ± 136 | 1,262 ± 139 | 1,330 ± 148 | 1,292 ± 143 | <0.001 |
| Pre-admission OAC | 123 (5.2) | 54 (10.6) | 90 (5.2) | 78 (6.8) | <0.001 |
| *1-year outcomes<sup>a</sup>* |  |  |  |  |  |
| Stroke recurrence<sup>b</sup> | 50 (2.1) | 14 (2.7) | 60 (3.5) | 34 (3.0) | 0.062 |
| Ischemic | 40 (1.7) | 6 (1.2) | 48 (2.8) | 26 (2.3) | 0.042 |
| Hemorrhagic | 2 (0.1) | 6 (1.2) | 5 (0.3) | 8 (0.7) | <0.001 |
| Death | 119 (5.0) | 50 (9.8) | 111 (6.5) | 115 (10.0) | <0.001 |

Values are n (%), mean ± SD, or median (IQR). P values from chi-square test for categorical variables and Kruskal–Wallis test for continuous variables. <sup>a</sup>Cumulative one-year outcomes include events recorded at both the 3-month and 1-year follow-up assessments. Follow-up was
available for 5,720 patients (99.7%). <sup>b</sup>In 17 patients, subtype was not available. Stroke etiology was classified according to the TOAST criteria: LAA, large-artery atherosclerosis; SVO, small-vessel occlusion; CE, cardioembolism; OD, other determined etiology; Undetermined includes two or more causes, negative evaluation, and incomplete evaluation. TOAST classification was unavailable in 505 patients (8.8%). CMB, cerebral microbleed; NIHSS, National Institutes of Health Stroke Scale; OAC, oral anticoagulant; WMH, white matter hyperintensity.

**Table 2.** Multivariable logistic regression for cerebral microbleed count ≥ 2, stratified by WMH burden.

|  | Full cohort |  | WMH-low |  | WMH-high |  |
| --- | --- | --- | --- | --- | --- | --- |
|  | OR (95% CI) | *P* | OR (95% CI) | *P* | OR (95% CI) | *P* |
| Atrial fibrillation | 1.04 (0.87–1.25) | 0.642 | 1.46 (1.11–1.93) | 0.007 | 0.95 (0.73–1.22) | 0.665 |
| Hypertension | 1.41 (1.23–1.60) | <0.001 | 1.11 (0.89–1.39) | 0.339 | 1.47 (1.24–1.75) | <0.001 |
| Interaction (AF × WMH-high) | OR 0.56 (0.41–0.77); *P* < 0.001 | | — | | | |

Models adjusted for age, sex, hypertension, diabetes mellitus, dyslipidemia, current smoking, coronary heart disease, and admission NIHSS. Only the primary exposure (atrial fibrillation) and the key contrasting predictor (hypertension) are shown. The interaction term was fitted in the full-cohort model. AF, atrial fibrillation; CI, confidence interval; OR, odds ratio; WMH, white matter hyperintensity.

### The four groups represent distinct clinical phenotypes

The four groups exhibited markedly different risk factor profiles (Table 1). Group 2 was distinguished by the highest prevalence of AF (28.0%) and cardioembolic etiology (26.7%), with coronary heart disease present in 11.4%; median WMH volume was 5.8 mL (IQR 3.6–9.0). Group 4 combined the highest hypertension prevalence (70.9%) with the highest lacune burden (median 3, 85.8% with at least one lacune). Group 1, the reference, had the lowest prevalence of all vascular risk factors. One-year stroke recurrence occurred in 158 patients (2.8%), with a pattern that differed qualitatively across groups (Table 1). In Groups 1, 3, and 4, ischemic recurrence predominated over hemorrhagic recurrence. In Group 2, ischemic and hemorrhagic recurrence occurred at identical rates (six raw hemorrhagic events, 1.2%; Aalen–Johansen 12-month cumulative incidence 0.96% with all-cause death as the competing event; Fig. 4a–c), a pattern unique to this group (hemorrhagic recurrence across groups, P < 0.001).

### AF prevalence increases with CMB count only in the low-WMH stratum

Within the WMH-low stratum, AF prevalence rose progressively with CMB count: 13.2% among patients with no CMBs, 17.9% with one, 27.1% with two to four, 32.9% with five to nine, and 28.6% with ten or more (P for trend < 0.001; Fig. 1b). No corresponding gradient was observed in the WMH-high stratum (P for trend = 0.73). The dose-response gradient for AF in the WMH-low stratum persisted after age adjustment (P = 0.008), whereas hypertension prevalence showed no age-adjusted gradient in either stratum (Fig. 1c; age-adjusted analysis in Supplementary Fig. 3).

### The AF–CMB association is confined to the low-WMH stratum

In the full cohort, AF was not associated with CMB count of two or more (OR 1.04, 95% CI 0.87–1.25; P = 0.642; Table 2). Stratification yielded divergent estimates: the WMH-low OR was 1.46 (95% CI 1.11–1.93; P = 0.007) and the WMH-high OR was 0.95 (95% CI 0.73–1.22; P = 0.665). Hypertension showed the inverse pattern, reaching significance in the WMH-high stratum (OR 1.47, 95% CI 1.24–1.75; P < 0.001) with a null estimate in the WMH-low stratum (OR 1.11, 95% CI 0.89–1.39; P = 0.339). The multiplicative interaction term yielded an OR of 0.56 (95% CI 0.41–0.77; P < 0.001; Table 2).

### The discordant group shows cortical predominance with deep structure sparing

Among 2,474 patients with at least one anatomically classified CMB, Groups 2 and 4 differed in spatial composition despite both carrying high CMB burden (Supplementary Table 2). Group 2 showed fronto-temporal cortical predominance with relative sparing of deep structures, whereas Group 4 had substantial involvement of both cortical and deep compartments. Under the Boston criteria version 2.0,^30^ 51.6% of Group 2 patients were classified as strictly lobar compared with 27.5% in Group 4 (Supplementary Table 2).

At equivalent total CMB counts, AF was associated with a lower proportion of deep CMBs (IRR 0.80, 95% CI 0.65–0.99; P = 0.040) without significant shifts in lobar or infratentorial proportions, and the effect did not differ between WMH strata (interaction P > 0.3 in all compartments; Supplementary Table 3).

Voxel-wise probability maps (Fig. 2) showed that Groups 2 and 4, despite sharing high CMB burden, differed in spatial architecture: cortical involvement was comparable in extent, but Group 2 showed conspicuous attenuation in deep gray matter structures that were densely involved in Group 4. Regional comparison normalized to total burden confirmed that this divergence was concentrated in the thalamus and putamen, where Group 4 showed marked excess, with no consistent cortical predominance in either direction (Fig. 3a). Voxel-wise permutation testing (5,000 permutations of group labels, family-wise-error-corrected at P < 0.05) confirmed 1,664 significant voxels, all localized to deep gray matter (thalamus, putamen, periventricular structures) and all in the direction of Group 4 excess; no voxel reached FWE significance for Group 2 excess (maximum positive t = 2.24; Fig. 3b). Region-level analysis with Benjamini-Hochberg FDR correction (Fig. 3c) ranked the thalamus as the most divergent region (mean 0.54 CMBs per patient in Group 2 vs 1.79 in Group 4; difference −1.25, 95% CI −1.50 to −1.01; FDR P < 0.001), followed by the middle temporal gyrus and putamen, and after burden normalization the top 12 regions split bidirectionally: deep and infratentorial structures (thalamus, putamen, cerebellum) showed Group 4 excess, whereas sensorimotor and limbic cortex (postcentral, frontal middle, cingulum mid, parietal inferior, calcarine, cuneus, precentral, occipital middle) showed Group 2 excess.

**Figure 2.**
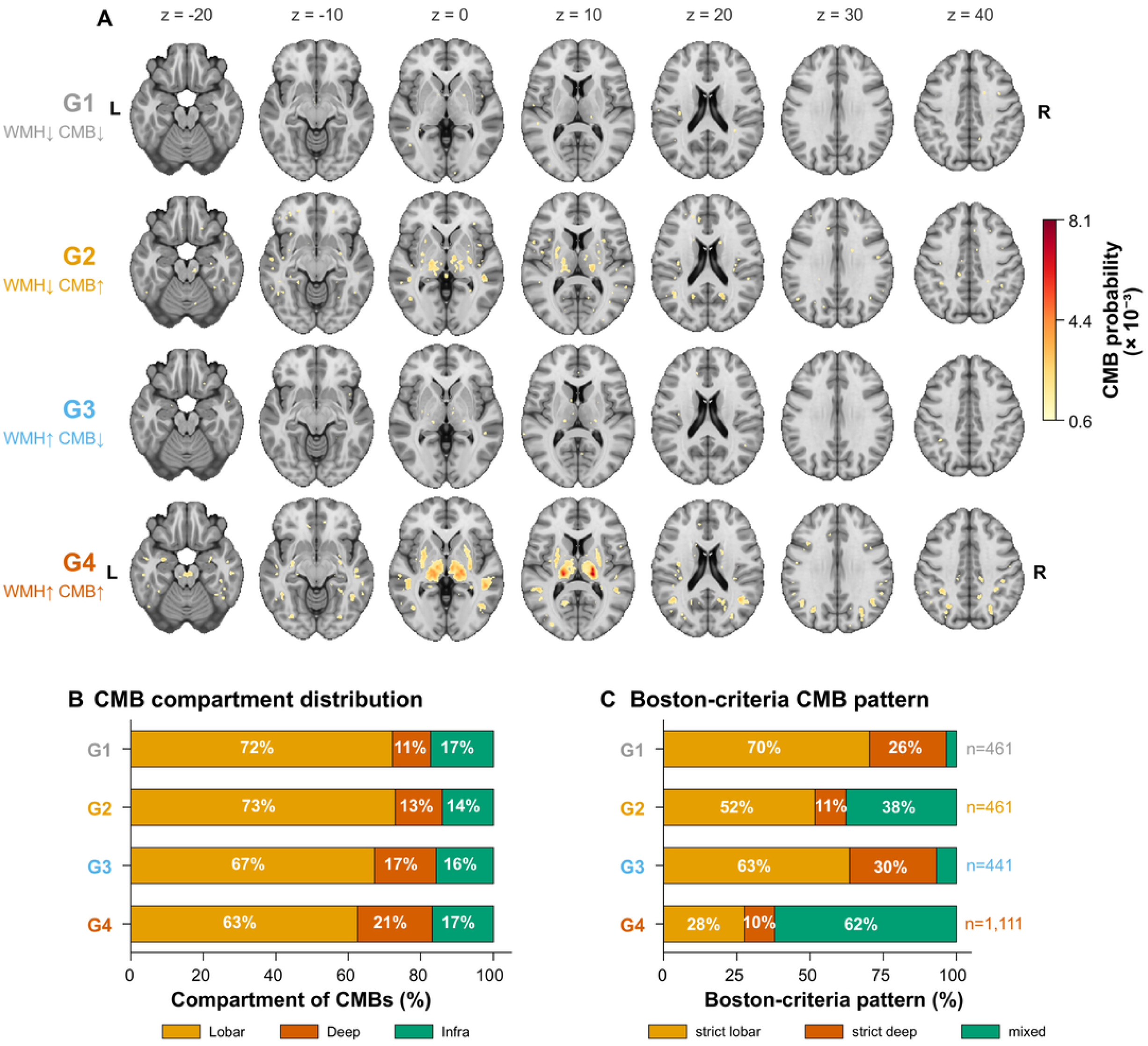
Cerebral microbleed spatial distribution differs systematically between WMH × CMB phenotypes. (a) Voxel-wise microbleed probability maps for each of the four phenotypic groups, overlaid on the MNI152 standard-space template (1 mm) and smoothed with a 3 mm FWHM Gaussian kernel. Seven axial slices are shown per group (z = −20, −10, 0, 10, 20, 30, 40 mm). Color intensity represents the per-voxel probability of microbleed presence, scaled to the global maximum across groups. Group 2 (WMH-low/CMB-high) shows fronto-temporal cortical predominance with attenuation in deep gray matter; Group 4 (WMH-high/CMB-high) shows additional dense involvement of the thalamus, putamen, and infratentorial structures. (b) Per-patient mean microbleed compartment fraction (lobar, deep, infratentorial), among microbleed-positive patients with anatomically classified lesions (n = 2,474). (c) Modified Boston criteria version 2.0 pattern by group (per-group n = 461 / 461 / 441 / 1,111 for G1 / G2 / G3 / G4), restricted to patients with at least one anatomically classified microbleed. Alt text: Three-panel figure. Panel a shows axial brain slices with voxel-wise microbleed probability heatmaps for each of the four phenotypic groups, demonstrating fronto-temporal cortical predominance in Group 2 and additional deep gray matter involvement in Group 4. Panel b is a stacked bar chart of mean per-patient microbleed compartment fractions (lobar, deep, infratentorial) by group. Panel c is a stacked bar chart showing the distribution of Boston criteria patterns (strict-lobar, mixed, strict deep) by group, with Group 2 dominated by the strict-lobar pattern and Group 4 dominated by the mixed pattern.

**Figure 3.**
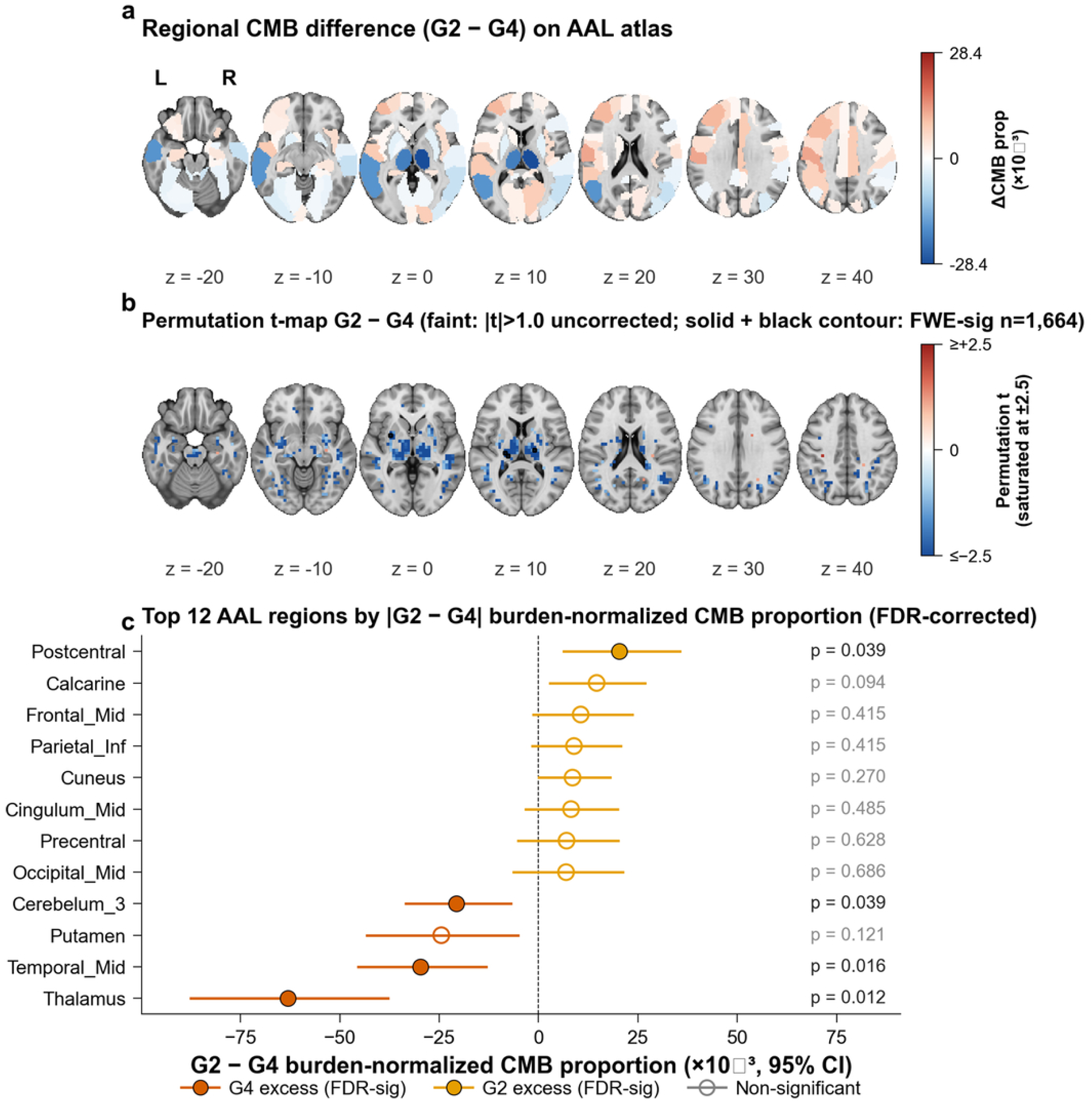
Voxel-wise and region-level spatial dissociation between Group 2 and Group 4. (a) Regional difference in per-microbleed proportion between Group 2 (WMH-low/CMB-high) and Group 4 (WMH-high/CMB-high), mapped onto the Automated Anatomical Labeling atlas. Each region is colored by the difference in normalized CMB proportion (Group 2 minus Group 4); red indicates relative Group 2 excess and blue indicates relative Group 4 excess. Deep structures including the thalamus and putamen show prominent Group 4 excess, whereas cortical regions show a mixed pattern without consistent Group 2 predominance. Axial slices at z = −20, −10, 0, 10, 20, 30, and 40 mm in MNI standard space. Regions outside the AAL parcellation (white matter, ventricles) display the underlying MNI152 template in grayscale. (b) Voxel-wise t-statistic map from permutation testing (5,000 permutations of group labels, family-wise error corrected at P < 0.05). Significant voxels are confined to deep gray matter (thalamus, putamen, periventricular structures), all in the direction of Group 4 excess; no voxel reached significance for Group 2 excess (maximum positive t = 2.24). FWHM 3 mm Gaussian smoothing applied. (c) Top 12 bilateral AAL regions ranked by absolute G2 − G4 difference in burden-normalized per-CMB proportion (each patient’s region count divided by their total CMB count, factoring out the higher absolute burden of Group 4). Markers indicate the mean per-mille difference; horizontal lines show 95% percentile bootstrap CIs (5,000 patient-level iterations); fill/open status reflects FDR-corrected bootstrap two-sided p-values from the same resampling distribution; filled circles, FDR-corrected P < 0.05; open circles, non-significance. Group 4 excess (vermillion) regions concentrate in deep and infratentorial structures (thalamus, mean per-mille difference −63; middle temporal gyrus; putamen; cerebellum 3); Group 2 excess (orange) regions occupy sensorimotor and limbic cortex (postcentral, frontal middle, cingulum mid, parietal inferior, calcarine, cuneus, precentral, occipital middle), demonstrating bidirectional topographic dissociation rather than a uniform burden gradient. AAL, Automated Anatomical Labeling; CMB, cerebral microbleed; FDR, false discovery rate; FWE, family-wise error; MNI, Montreal Neurological Institute; WMH, white matter hyperintensity. Alt text: Three-panel figure comparing Group 2 and Group 4 microbleed spatial distributions. Panel a is a brain map showing regional differences in normalized microbleed proportion overlaid on axial slices, with deep structures in blue (Group 4 excess) and cortical regions showing mixed patterns. Panel b is a thresholded voxel-wise t-statistic map showing family-wise-error-corrected significance confined to deep gray matter structures, all in the direction of Group 4 excess. Panel c is a horizontal forest plot of region-level mean microbleed differences across Automated Anatomical Labeling regions, ranked by effect size, with the thalamus as the most divergent region.

Region-level enrichment analysis, computed as the ratio of observed CMB proportion to the proportion expected from each AAL region’s volume, confirmed that the spatial divergence between Groups 2 and 4 was driven by thalamic concentration in Group 4 (enrichment 14.1× relative to regional volume vs 9.9× in Group 2, P < 0.001). By volume-normalized enrichment, Group 2 showed relative concentration in the hippocampus (3.5× vs 2.5× in Group 4), insula, and mid-cingulate (Supplementary Fig. 4); the complementary burden-normalized analysis (Fig. 3c) ranked sensorimotor and limbic cortex (postcentral, frontal middle, parietal inferior, cingulum mid, calcarine, cuneus, precentral, occipital middle) as the regions of relative Group 2 excess, with the only voxel-wise FWE-significant findings localized to the deep gray matter direction of Group 4 excess.

### AF amplifies the spatial phenotype but does not originate it

Among 461 Group 2 patients with at least one anatomically classified CMB (matching the Figure 2 cohort), those with AF (n = 130) had fewer deep CMBs per patient than those without (mean 0.52 vs 0.81; P = 0.02 by Mann-Whitney; Supplementary Table 4). No anterior–posterior asymmetry was observed: the per-patient anterior ratio (frontal+temporal / frontal+parietal+temporal+occipital) did not differ between AF-positive and AF-negative subgroups (0.58 vs 0.58; P = 0.96). The topographic divergence between Groups 2 and 4 persisted among AF-negative patients (P < 0.001; Supplementary Table 4), confirming that AF status alone does not account for the spatial phenotype.

### Assessment of potential confounders

Three prespecified analyses addressed alternative explanations: pre-stroke antithrombotic use (Supplementary Tables 5, 7), AF reclassification by exposure duration (no AF / new-onset / prior; Supplementary Table 6), and collider-bias assessment of WMH residuals between AF-positive and AF-negative patients (Supplementary Fig. 5). All supported the primary stratum-specific finding.

### Sensitivity analyses

The interaction between AF and WMH stratum persisted across all twelve prespecified sensitivity analyses (Supplementary Table 7). Varying the CMB threshold, three alternative WMH definitions (raw volume split and two tertile splits), exclusion of cardioembolic strokes (n = 4,837), age-stratified subgroups (<65, 65–74, ≥75), additional adjustment for lacune count and total brain volume, and generalized estimating equations accounting for within-site clustering all preserved the WMH-low AF–CMB association (OR range 1.31–2.20) with null effects in the WMH-high stratum.

Treating WMH burden as continuous yielded a parallel pattern. The AF × WMH residual interaction was OR 0.65 (95% CI 0.52–0.82; P < 0.001), and the implied AF odds ratio for CMB declined monotonically across the WMH residual distribution, from 1.94 (1.42–2.67) at the 5th percentile to 1.27 (1.05–1.54) at the median and 0.72 (0.52–1.00) at the 95th percentile (Fig. 1d).

Hemispheric laterality analysis among Group 2 AF-positive patients with unilateral supratentorial infarcts did not support an acute-embolic origin for these CMBs (Supplementary Fig. 6).

### Hemorrhagic recurrence by phenotype

Despite the smallest sample size (n = 510), Group 2 carried disproportionate hemorrhagic risk (Fig. 4). In adjusted Firth penalized logistic regression, Group 2 had the highest odds of one-year hemorrhagic stroke recurrence among the four groups (Fig. 4a). On Aalen–Johansen analysis with all-cause death as the competing event, the 12-month cumulative incidence of hemorrhagic recurrence was 0.96% in Group 2, exceeding Group 4 (0.46%), Group 3 (0.15%), and Group 1 (0.00%; Fig. 4b, c). The contrast was specific to hemorrhagic recurrence: Group 2 was not the worst group on adjusted three-month or one-year mRS, ordinal mRS, or all-cause mortality, although directional point estimates favored Groups 1 and 3 (Fig. 4a). Cause-specific and Fine–Gray subdistribution hazard ratios for Group 2 vs Group 1 were concordant in direction, arguing against differential mortality alone as the driver of the elevated CIF estimate.

**Figure 4.**
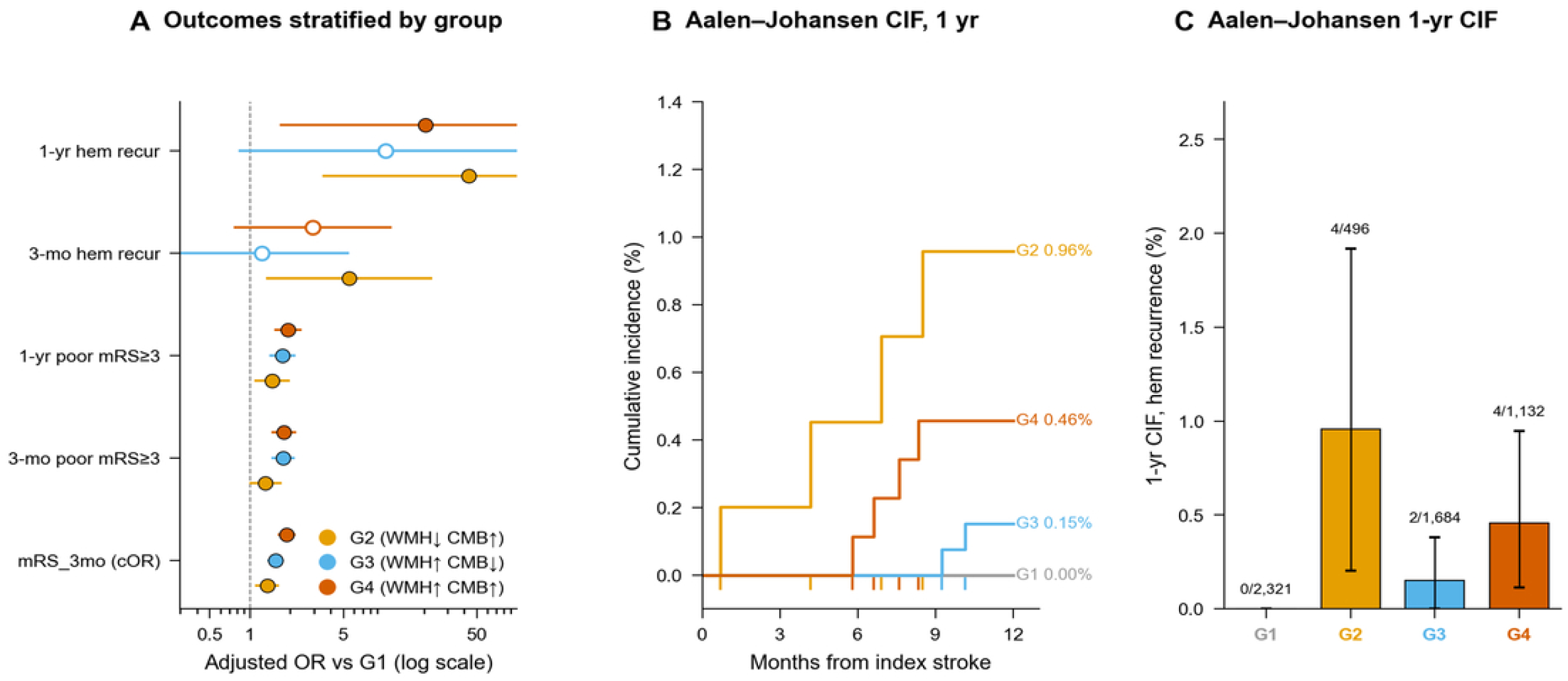
Spontaneous hemorrhagic recurrence is concentrated in the discordant WMH × CMB phenotype. (a) Adjusted ORs (95% CI) for spontaneous outcomes vs G1 (n = 5,720); filled circle = P < 0.05, open = NS. Firth logistic for hemorrhagic events; standard logistic for poor mRS; ordinal logistic for the cOR. All models adjusted for age, sex, log NIHSS, hypertension, diabetes, dyslipidemia, atrial fibrillation, TOAST. (b) Aalen–Johansen 12-month cumulative incidence of hemorrhagic recurrence by group, with all-cause death as the competing event. (c) The same CIFs at the 1-year endpoint shown as bars with 95% logit-CIs. *Abbreviations*: CI, confidence interval; CIF, cumulative incidence function; CMB, cerebral microbleeds; cOR, common odds ratio; HR, hazard ratio; mRS, modified Rankin Scale; NIHSS, National Institutes of Health Stroke Scale; NS, non-significant; OR, odds ratio; TOAST, Trial of Org 10172 in Acute Stroke Treatment; WMH, white matter hyperintensities. Alt text: Three-panel composite on hemorrhagic bleeding stratified by WMH × CMB phenotype. Panel a is a forest plot of adjusted odds ratios (log-scaled x-axis) for spontaneous outcomes versus Group 1, showing markedly elevated 1-year hemorrhagic recurrence and ordinal mRS odds for Group 2 and Group 4. Panel b shows Aalen–Johansen step curves of cumulative hemorrhagic recurrence over twelve months by group: Group 2 (orange) reaches the highest 0.96% with earliest events around months 1 and 4, Group 4 (vermillion) reaches 0.46% with events from month 6 onward, Group 3 (sky blue) ends at 0.15%, and Group 1 (gray) remains flat at 0%. Colored rug ticks below the x-axis mark individual event times. Panel c is a bar chart of the same 1-year cumulative incidences with 95% confidence intervals, preserving the Group 2 > Group 4 > Group 3 > Group 1 ordering.

## Discussion

The principal finding of this study is that AF is independently associated with CMBs only when WMH burden is low relative to age, and that this association is absent in the high-burden stratum. What appears in the unstratified cohort as a null effect (OR 1.04, P = 0.642) is the arithmetic residue of two divergent signals (WMH-low OR 1.46, WMH-high OR 0.95) whose cancellation the interaction term formally registers (OR 0.56, P < 0.001). That prior studies have yielded inconsistent AF–CMB associations is, in this light, a predictable consequence of pooling patients in whom the two signals offset one another.

These findings are consistent with two distinct vascular contexts for CMB formation. Where WMH burden is high, CMBs conform to the hypertensive phenotype with deep and infratentorial predominance. Where burden is low, CMBs concentrate in the frontal and temporal cortex with conspicuous sparing of deep structures, and AF emerges as the significant predictor. The offset model indicates that AF does not drive CMBs toward the cortex but away from deep structures. AF does not, however, appear to give rise to this spatial phenotype on its own: the topographic divergence persists among patients without the arrhythmia, and new-onset and prior AF confer indistinguishable risk. One interpretation is that AF and lobar-predominant CMBs may share an upstream vascular substrate that affects both the cortical microvasculature and the atrial myocardium, although this remains to be demonstrated mechanistically.

The inconsistency of prior AF–CMB associations can now be understood as unrecognized effect modification. Studies pooling patients across the full spectrum of white matter disease would be expected to detect only a diluted signal: the Microbleeds International Collaborative Network reported modest associations across heterogeneous cohorts,^4^ and the Swiss-AF study found CMB prevalence no higher in AF patients than in controls despite greater WMH burden.^31^ Our interaction model renders both findings intelligible, as the AF–CMB association strong in the low-WMH stratum is nullified in the high-WMH stratum, producing attenuation or reversal when the two are combined. A complementary line of evidence comes from spatiotemporal analysis of WMH progression, which identified a temporo-occipital WMH subtype specifically enriched for AF.^32^ While that work characterized the white matter landscape of AF, the present study characterizes the CMB landscape that emerges in its relative absence. Together, these findings suggest that the cerebrovascular signature of AF may be distinct from that of hypertensive arteriopathy and may be obscured whenever composite small vessel disease metrics treat these pathways as one.

The biological substrate linking AF to lobar-predominant CMBs in a low-WMH context remains incompletely resolved. Cerebral amyloid angiopathy is the most intuitive candidate,^33^ and amyloid co-pathology in this group cannot be excluded without APOE genotyping or cortical superficial siderosis data. Several features, however, resist a straightforward amyloid interpretation: the cortical distribution showed fronto-temporal rather than posterior predominance,^34^ and the AF association was strongest in patients younger than 65 and attenuated with age, the opposite of the gradient expected if age-dependent amyloid accumulation were the driving mechanism. A thromboembolic account is similarly difficult to sustain, as the association persisted after exclusion of all cardioembolic strokes. An alternative possibility, grounded in the concept of atrial cardiomyopathy,^35^ is that AF and cortical microvascular fragility may be joint manifestations of a diffuse vascular process; this would be consistent with the equal effect sizes of new-onset and prior AF, the persistence of the spatial phenotype without the arrhythmia, and the null effect of anticoagulant use, but remains an untested hypothesis. Regardless of the ultimate mechanism, this phenotype may be overlooked by WMH-based assessment and warrants independent recognition.

These observations carry a direct implication for how cerebral small vessel disease is assessed in clinical practice. WMH volume, whether measured quantitatively or graded visually, is the most widely used index of subclinical cerebrovascular injury and figures prominently in risk stratification for anticoagulation, cognitive decline, and surgical decision-making.^36,37^ Yet nearly one in ten patients in this cohort harbored a CMB burden disproportionate to their WMH load, a phenotype invisible to evaluation that begins and ends with white matter. That this group was enriched for AF, carried a distinct spatial signature, and exhibited risk factor profiles unlike those of patients with concordantly elevated markers suggests that WMH and CMB count may index partially distinct vascular processes, such that WMH burden alone may not capture the full topography of small vessel injury. Incorporating CMB presence and distribution into the clinical interpretation of brain MRI, particularly in patients with AF and unexpectedly low WMH burden, may identify a vascular phenotype that conventional assessment frameworks do not address.

Outcome data illustrate the clinical stakes of this phenotype. In Groups 1, 3, and 4, ischemic stroke recurrence outnumbered hemorrhagic recurrence by ratios of 20:1, 10:1, and 3:1, consistent with the ischemic predominance that underpins anticoagulation guidelines.^1,2^ Group 2 was the sole exception: ischemic and hemorrhagic recurrences occurred at identical rates (six events each, 1.2%). The small number of hemorrhagic events precludes formal inference and these observations should be regarded as hypothesis-generating, but the parity is notable in a group whose high AF prevalence typically leads to anticoagulation at discharge and whose defining cortical microvasculature carries an a priori bleeding tendency. The CROMIS-2 cohort has shown that increasing CMB burden progressively narrows the margin between ischemic benefit and hemorrhagic risk in anticoagulated AF patients.^1^ Our descriptive data raise the possibility that this margin may be reached at comparatively modest CMB counts when the underlying vasculopathy is predominantly cortical, since Group 4 maintained a 3:1 ischemic-to-hemorrhagic ratio despite a higher median CMB burden. These findings do not argue against anticoagulation when otherwise indicated,^38^ but they suggest that risk stratification weighing CMB count without reference to WMH context may warrant prospective evaluation in this specific phenotype.

Several limitations warrant explicit acknowledgment. The cross-sectional design precludes causal inference, though the E-value of 2.29 for the primary estimate (1.46 for the lower confidence bound) indicates that an unmeasured confounder would need to be associated with both AF and CMB burden more strongly than any known candidate, including APOE ε4, to explain away the observed association.^29^ The absence of APOE genotyping and cortical superficial siderosis data prevents direct adjudication of the amyloid angiopathy hypothesis. AF was treated as a binary exposure in the primary model, although new-onset and prior AF were examined as a sensitivity analysis (Supplementary Table 6); information on AF subtype (paroxysmal/persistent/permanent), duration, and left atrial morphology was not available in the registry. The 4,191 excluded patients were enriched for AF (28.6%) and cardioembolic etiology (27.6%), with higher median admission NIHSS (Supplementary Table 1), rendering the analytic cohort a conservative subsample in which the true AF–CMB association is likely attenuated. The spatial analysis excluded 5.9% of individual CMBs whose centroids fell outside AAL atlas-defined regions, predominantly those located in white matter or at the interface between parcellated structures. The cohort is drawn from a single East Asian population, and generalizability to other ethnicities remains untested. Against these constraints stand several methodological assets, including quantitative, algorithm-derived imaging markers applied uniformly across all patients, voxel-level spatial data in over 2,500 individuals, formal interaction testing with twelve concordant sensitivity analyses, and systematic evaluation of collider bias, antithrombotic exposure, and unmeasured confounding through the E-value.

## Conclusions

In a large multicenter cohort of acute ischemic stroke patients with quantitative MRI-derived small vessel disease markers, the association between AF and CMBs was confined to patients whose WMH burden was low relative to age. This context-dependent association was accompanied by a topographically distinct CMB distribution characterized by deep-structure sparing and was not attributable to anticoagulant use. These findings suggest that the vascular interpretation of CMBs is context-dependent, with the WMH milieu shaping the likely underlying pathology, and that clinical assessment of cerebral small vessel disease based on WMH alone may overlook a CMB phenotype linked to AF.

## Data availability

Anonymized data supporting the findings of this study are available from the corresponding author upon reasonable request, subject to institutional data-sharing agreements and privacy regulations. The deep learning segmentation algorithms used to extract white matter hyperintensity volume and cerebral microbleed count are described in reference 16 and are available for non-commercial research use upon reasonable request to the corresponding author, subject to institutional data-sharing policies.

## Acknowledgments

We thank the research coordinators and stroke nurses at all participating centers for their contributions to data collection.

## Author contributions

All authors contributed to the study concept and design, interpretation of results, and critical revision of the manuscript. W.S.R. led the conceptualization, methodology development, and supervision of the imaging analysis pipeline, including the segmentation algorithms and quantitative biomarker extraction. M.J.L. contributed to the development and validation of the imaging algorithms. L.S. provided neuroradiological interpretation and image quality control. The statistical analysis of clinical outcomes and the multivariable modeling were performed independently by B.J.K., who had full access to the deidentified analytic dataset. W.S.R. and B.J.K. drafted the original manuscript and prepared the data visualizations. B.J.K. led the supervision, funding acquisition, and project administration. K.K., J.Gk.K., S.J.L., J.K.C., T.H.P., J.Y.L., K.L., D.H.K., J.L., H.K.P., Y.J.C., K.S.H., M.L., M.S.O., K.H.Y., D.S.G., D.E.K., H.K., J.T.K., J.Jg.K., J.C.C., W.J.K., J.H.K., K.S.Y., D.I.S., J.H.H., S.I.S., S.H.L., C.K., H.B.J., K.Y.P., K.J.L., C.K.K., J.K., J.Y.K., and H.J.B. contributed to data acquisition and curation. All authors had full access to the data, reviewed the final manuscript, and approved it for submission.

## Supplementary material

Supplementary material is available at Stroke online.

